# Rethinking Input Complexity in Transformer-Based Clinical Prediction: Implications for Feature Dimensionality and Sequence Length in Longitudinal EHR Data

**DOI:** 10.64898/2026.09.03.26362165

**Authors:** Wansu Chen, Botao Zhou, Robert S. Zeiger, William Crawford, Michael Schatz, Eric Puttock, Stanley Xu, Matthew T. Slaughter, Fagen Xie

## Abstract

**Objective:** Transformer-based models for clinical prediction using longitudinal electronic health record (EHR) data are often developed with large feature sets and long patient histories under the assumption that more data improves performance. However, high-dimensional inputs and long sequences increase computational burden, potentially limiting scalability. We evaluated how feature dimensionality and sequence length affect predictive performance, calibration, risk stratification, and computational efficiency in EHR prediction.

**Methods:** Using longitudinal EHR data from adults with mild asthma in an integrated healthcare system, we evaluated input representation design for predicting acute asthma exacerbation. Feature dimensionality was reduced using Integrated Gradients attribution scores, univariate performance-based selection, and clinically guided selection strategies. Sequence length was varied using percentile-based truncation of patient histories. Performance was assessed using discrimination, calibration, high-risk classification, threshold-based event capture, and computational efficiency across regions.

**Results:** Models using fewer features achieved discrimination comparable to the 80-feature reference model, with AUROC values ranging from 0.843 to 0.864 versus 0.870 for the full model. Moderate sequence-length truncation reduced training time by more than 70% with minimal loss in discrimination. Although reduced-dimensional models showed attenuation of predicted risk at the upper tail, they identified similar high-risk populations and captured comparable proportions of asthma exacerbation events at clinically relevant thresholds.

**Conclusion:** Transformer-based prediction models maintained strong performance across reduced feature sets. While dimensionality reduction modestly affected calibration at the highest risk levels, moderate sequence-length reduction substantially reduced computational burden with limited change in overall discrimination. These findings highlight trade-offs between input complexity, predictive performance, and computational efficiency.

**Funding:** National Heart, Lung, and Blood Institute, National Institutes of Health (R01 HL163049).

**Lay Summary:** Artificial intelligence models are increasingly used to predict clinical outcomes from electronic health record (EHR) data. Many modern deep learning models are built using large numbers of clinical variables and long patient histories because more data is often assumed to improve prediction. However, these complex models require substantial computing resources and may be difficult to implement in real-world healthcare systems.

In this study, we examined whether simpler input designs could maintain strong predictive performance while reducing computational burden. Using transformer-based models and longitudinal EHR data from adults with mild asthma, we systematically evaluated how the number of clinical features and the length of patient histories affected prediction performance and efficiency.

We found that models using fewer features and shorter patient histories performed similarly to more complex models for identifying patients at high risk of acute asthma exacerbation. Although simplified models showed modest differences in predicted risk estimates at the highest risk levels, they identified similar high-risk patient groups and captured similar numbers of asthma exacerbation events.

These findings suggest that deep learning models for healthcare prediction may not require highly complex input designs to achieve clinically useful performance, which could improve scalability and real-world implementation.

## INTRODUCTION

Deep learning models are increasingly used for clinical risk prediction using longitudinal electronic health record (EHR) data. [1–3] Transformer-based architectures, in particular, enable modeling of temporally ordered clinical events and have demonstrated strong performance across a range of prediction tasks by capturing complex dependencies within patient histories. As a result, there has been growing interest in applying transformer-based models to large-scale EHR data to support risk stratification and population-level clinical decision-making. [4–7]

In practice, these models are often developed under the implicit assumption that more data leads to better performance, including the use of large feature sets and long longitudinal histories. However, high-dimensional input representations introduce substantial practical challenges. Large numbers of features increase data engineering complexity and require stable availability of heterogeneous data elements across sites, time periods, and clinical workflows, which may limit model portability and robustness after deployment. [8, 9] Similarly, long patient histories increase computational cost for both training and inference, particularly for transformer-based models where complexity scales with sequence length. [10, 11] These factors can hinder scalability, delay model updates, and create barriers to routine use in real-world health systems.

Despite these challenges, there is limited empirical guidance on how input design choices, specifically feature dimensionality and sequence length, affect clinically relevant model behavior. Prior work in traditional machine learning has emphasized dimensionality reduction to improve generalizability and interpretability, whereas deep learning approaches often rely on representation learning without explicit feature selection. [12, 13] In the context of EHR-based modeling, studies examining dimensionality reduction, sequence compression, or truncation strategies have primarily emphasized optimization-oriented metrics such as training loss and discrimination, whereas comparatively less attention has been devoted to calibration, clinical utility, and threshold-based identification of high-risk patients. [14–16] As a result, it remains unclear how much input complexity is necessary to achieve robust and clinically useful performance.

In this study, we address this gap by systematically evaluating feature dimensionality and sequence length as key design parameters in transformer-based clinical risk prediction using longitudinal EHR data. Using acute asthma exacerbation in adults with mild asthma as a representative use case, we compare multiple feature selection strategies and sequence-length configurations under controlled conditions while holding model architecture constant. We assess the impact of these design choices on discrimination, calibration, risk stratification, threshold-based event capture, and computational efficiency. By providing a comprehensive evaluation of input representation trade-offs, this work aims to inform the development of scalable and deployable deep learning models for clinical prediction in real-world healthcare settings.

## METHODS

### Study Overview

We conducted a retrospective methodological evaluation to examine how input dimensionality influences predictive performance and computational efficiency in deep learning–based clinical risk prediction models. Acute asthma exacerbation in patients with mild asthma was used as a representative use case within a previously established EHR cohort of adults with mild asthma. [17, 18]

The study design held the study population, outcome definition, temporal encoding of EHR data, and transformer architecture constant while systematically varying feature dimensionality, feature-selection strategy, and maximum sequence length (Figure 1). Experiments were conducted under controlled conditions to isolate the effects of these input representation design choices on discrimination, calibration, risk stratification, threshold-based event capture, and computational efficiency.

**Figure 1.** Study design and experiment framework. Study design overview illustrating the evaluation of input dimensionality in transformer-based clinical risk prediction using longitudinal electronic health record (EHR) data. The study population and data framework, longitudinal sequence representation, model architecture, and training procedures were held constant across all experiments. Two experimental axes were evaluated. In feature dimensionality experiments, feature selection strategy (Integrated Gradients, univariate AUC-based, and clinically guided) and feature set size were varied under a fixed maximum sequence length, with model performance assessed using discrimination, calibration, risk stratification, threshold-based event capture, and computational efficiency. In sequence-length experiments, the maximum input length was varied using percentile-based truncation under a fixed 20-feature Integrated Gradients–selected model, with evaluation focused on discrimination and computational efficiency.

### Data Source, Study Population, and Outcome

The analytic dataset was derived from structured EHR data from Kaiser Permanente Southern California (KPSC), including diagnoses, medications, laboratory results, and healthcare encounters.

Cases were defined as adults aged 18–85 years who experienced an AAE during 2013–2019, with the AAE date serving as the index date. Eligible cases required at least one asthma diagnosis and ≥12 months of continuous enrollment before the index date and met criteria for mild asthma, defined by low-intensity controller therapy, limited short-acting beta-agonist use, and absence of markers of moderate-to-severe disease, including asthma-related hospitalization and selected comorbid conditions in the past 12 months. Patients could contribute multiple eligible case events during the study period.

Controls were selected from eligible patients without AAE during the study period, and assigned case-derived index dates to align predictor assessment windows. Each control patient contributed only one observation to the analytic dataset.

### Outcome Definition

AAE was defined using structured EHR criteria including asthma-related hospitalization, emergency department or observation encounters, or systemic corticosteroid treatment associated with asthma-related diagnoses.

### Baseline Transformer Architecture and Longitudinal Input Encoding

Predictor variables were ascertained from the 24 months preceding the index date.

Structured EHR data were transformed into temporally ordered longitudinal sequences representing clinical events occurring during the observation window (Supplementary Figure E1). Each clinical concept (e.g., diagnosis, medication, laboratory result, or healthcare encounter) was represented as an individual token, with multiple tokens permitted on the same calendar day. Demographic characteristics were encoded as static patient-level variables. Clinical events were arranged chronologically to form the longitudinal input sequence for transformer-based modeling.

The transformer-based architecture and training framework were held constant across all experiments. Model inputs consisted of embedded longitudinal token sequences with positional encoding to preserve temporal order.

Training procedures and hyperparameters were held constant across experiments to isolate the effects of feature dimensionality and sequence length.

### Dimensionality Reduction Strategies and Feature Subsets

To evaluate the effect of dimensionality reduction, we applied three complementary feature-selection strategies to the 80-feature baseline input space (Table 1). For all experiments, reduced-dimensional models were trained using the same transformer architecture and training settings as the full-feature reference model, with only the input feature set modified. For the two data-driven strategies, feature rankings were derived once from the full dataset to define fixed feature subsets that were applied consistently across all cross-validation experiments, thereby enabling direct comparison of identical input representations across model configurations.

**Table 1.** Selection Strategies Evaluated for Dimensionality Reduction in Transformer-Based Risk Prediction.

| Method | Type | Feature Input | Selection Basis | Feature Subsets Tested | Description |
| --- | --- | --- | --- | --- | --- |
| Clinically guided | Expert-derived | 80 | Clinical relevance and guidelines | 20 | Benchmark for interpretability and feasibility |
| Integrated Gradients (IG) | Model-based (deep learning) | 80 | Mean absolute attribution score | 10, 20, 40 | Captures non-linear feature importance from the trained model |
| AUC-based selection | Univariate model-based | 80 | Individual feature discrimination (AUC) | 10, 20, 40 | Ranks features by their standalone AUC value |

Feature counts shown under “Feature Input” reflect the full baseline structured feature space available prior to selection. Reduced feature subsets were derived exclusively from the training data and then fixed and applied identically for all subsequent model training and evaluation. Clinically guided features were selected a priori based on clinical expertise, guideline-informed reasoning, and prior asthma risk prediction studies, and did not rely on data-driven ranking. All feature selection experiments were conducted under a fixed maximum sequence length unless otherwise specified.

Integrated Gradients (IG)–based feature selection quantified each feature’s contribution to model output using attribution scores derived from the trained reference transformer model. IG is a gradient-based attribution method that estimates feature importance by integrating gradients from a baseline input to the observed input and has been widely used to interpret deep neural network predictions. [19, 20] Attribution values were computed across the training set and summarized using the mean absolute IG score for each feature. [19] Features were then ranked by importance, and reduced feature subsets were defined using the top 10, top 20, and top 40 ranked features. Features included in each IG-derived subset are listed in Table E1.

Univariate AUC-based feature selection ranked features according to their individual discriminatory ability for predicting acute asthma exacerbation. Each feature was evaluated independently using its area under the receiver operating characteristic curve (AUROC) within the training data. Features were subsequently ranked by AUROC, and reduced feature subsets were defined using the top 10, top 20, and top 40 ranked features. Features included in each AUC-derived subset are listed in Table E1.

Clinically guided feature selection used a fixed subset of 20 features curated a priori based on clinical expertise and guideline-informed reasoning (Table E1). [13] This subset was intended to reflect clinically interpretable variables commonly available in real-world healthcare settings. Feature selection was informed by prior evidence on risk factors for acute asthma exacerbation, including findings from our previously published work. [18]

### Sequence-Length and Runtime Design

To evaluate the impact of maximum sequence length on predictive performance and computational efficiency, we systematically varied the maximum sequence length (max_len) applied to longitudinal input sequences while holding all other model components constant.

A maximum sequence length of 512 tokens was selected as the reference configuration based on the empirical distribution of sequence lengths (Figure 2). This threshold retained most longitudinal histories while limiting excessive computational burden associated with very long sequences. Sequences exceeding this length were truncated to retain the most recent tokens, whereas shorter sequences were padded.

**Figure 2.** Effect of maximum sequence length on training time and discrimination. Mean training time (left y-axis, blue) and AUROC (right y-axis, red) are shown as a function of maximum sequence length, evaluated using the Integrated Gradients–selected 20-feature model. Shaded regions indicate variability across cross-validation folds. Reducing maximum sequence length was associated with substantial decreases in training time, while AUROC remained largely stable across moderate sequence-length settings, with more pronounced performance degradation observed only at the most restrictive settings.

Alternative maximum sequence length settings were defined using percentile-based cutoffs derived from the empirical sequence-length distribution, including the 98th, 95th, 90th, 85th, 80th, 75th, 70th, 65th, 60th, 55th, and 50th percentiles. For each percentile threshold, max_len was defined such that the corresponding proportion of patient sequences could be retained without truncation, while longer sequences were truncated to preserve the most recent longitudinal information.

Because sequence lengths demonstrated a pronounced right-skewed distribution, percentile-based truncation enabled systematic evaluation of trade-offs between retained longitudinal information and computational efficiency. Evaluation was limited to the 50th percentile to avoid excessive truncation of patient histories.

All sequence-length experiments were conducted using the IG-selected 20-feature model. Feature-selection strategy, feature count, transformer architecture, and training procedures were held constant to isolate the independent effect of sequence length. Batch size was fixed at 32 across experiments.

### Model Training and Evaluation

#### Cross-site validation design

Model development and evaluation were conducted using six-fold cross-validation defined by geographically distinct Medical Service Areas (MSAs). In each iteration, data from five MSAs were used for training and the remaining MSA was held out for evaluation. Performance metrics were averaged across folds. This design approximates an internal–external validation framework by evaluating model performance across geographic and operational variation. [21]

#### Discrimination Performance

Model discrimination was assessed using the area under the AUROC and the area under the precision–recall curve (AUPRC) [14], calculated on held-out evaluation data within each cross-validation fold.

#### Calibration and Risk Stratification Across Feature Set

Calibration and risk stratification analyses evaluated how feature dimensionality influenced predicted risk distributions, patient-level risk assignment, and high-risk identification. Shared risk bins derived from the full-feature reference model were applied unchanged across all reduced-feature models. Mean predicted risk and observed event rates were calculated within each bin to assess calibration across the risk distribution (Figure 3A).

**Figure 3.** Distribution of observed sequence lengths and percentile-based maximum sequence length settings. Histogram showing the empirical distribution of observed sequence lengths across the study cohort, where sequence length is defined as the number of structured EHR input elements included in a longitudinal observation. Vertical dashed lines indicate percentile-based maximum sequence length settings evaluated in subsequent analyses, ranging from the 98th percentile (p98) to the 50th percentile (p50). The pronounced right-skew of the distribution indicates that a small fraction of observations have substantially longer sequences than the majority, motivating evaluation of alternative maximum sequence length configurations to improve computational efficiency while preserving predictive performance. The vertical line at 512 indicates the selected maximum sequence length used in model training, chosen to balance sequence coverage and computational efficiency.

To evaluate patient-level risk assignment, predicted risks from reduced-feature models were compared against predictions from the full-feature reference model among patients with predicted risk ≥0.8 under the reference model (Figure 3B).

For operational risk stratification analyses, a fixed predicted-risk threshold derived from the reference model was used to classify approximately 20% of patients as high risk (Figure 3C). Event capture was assessed as the proportion of observed exacerbations occurring among patients classified as high risk (Figure 3D).

#### Computational Efficiency

Computational efficiency was quantified using total training time, inference time per patient record, and throughput, defined as the number of patient records processed per second. Runtime comparisons were conducted under standardized hardware and software conditions.

### Sensitivity Analysis

Sensitivity analyses were conducted using batch sizes of 32, 64, and 128 while holding all other model parameters constant. Discrimination and runtime metrics were evaluated across cross-validation folds.

Sensitivity of risk stratification to selection thresholds was additionally evaluated using cumulative event capture curves. Patients were ranked by predicted risk, and the cumulative proportion of observed events captured was calculated as progressively larger fractions of the population were selected (Figure E2). This analysis complements fixed-threshold evaluation across broader population strata.

### Software

All analyses were performed in Python. Deep learning models were implemented using PyTorch and the Hugging Face Transformers library. Data preprocessing and evaluation were conducted using NumPy, pandas, and scikit-learn.

### Ethics Approval

This study was approved by the Kaiser Permanente Southern California Institutional Review Board with a waiver of informed consent for retrospective analysis.

## RESULTS

### Patient Characteristics

A total of 395,941 observations were included in the analytic dataset, comprising 86,424 cases with AAE and 309,517 controls without. These observations correspond to eligible index events, and individual patients could contribute multiple events. The mean (±SD) age was 44.7 (±17.9) years, and 63.9% were female.

Baseline characteristics stratified by case status are summarized in Table 2. Compared with controls, cases were older (49.2 vs 43.4 years) and more likely to be female (72.4% vs 61.6%). Indicators of asthma-related disease activity and treatment intensity were consistently more prevalent among cases, including higher use of short-acting β-agonists (87.7% vs 72.2%), asthma controller medications (66.3% vs 38.9%), combination ICS/LABA therapy (22.6% vs 9.6%), and systemic corticosteroids (72.7% vs 38.5%).

**Table 2.** Patient Characteristics.

| **Characteristics** | **Overall**<br>**(N = 395,941)** | **Cases**<br>**(N = 86,424)** | **Controls**<br>**(N = 309,517)** |
| --- | --- | --- | --- |
| **Demographics** |  |  |  |
| Age, mean (SD) | 44.7 (17.9) | 49.2 (16.7) | 43.4 (18.0) |
| Female, n (%) | 253,042 (63.9) | 62,535 (72.4) | 190,507 (61.6) |
| **Asthma-related characteristics** |  |  |  |
| Short-acting β-agonist use, n (%) | 299,337 (75.6) | 75,770 (87.7) | 223,567 (72.2) |
| Asthma controller use, n (%) | 177,811 (44.9) | 57,309 (66.3) | 120,502 (38.9) |
| ICS/LABA use, n (%) | 49,274 (12.4) | 19,505 (22.6) | 29,769 (9.6) |
| Systemic corticosteroid use, n (%) | 181,832 (45.9) | 62,795 (72.7) | 119,037 (38.5) |
| **Comorbidities / phenotype** |  |  |  |
| Allergic rhinitis, n (%) | 88,674 (22.4) | 24,133 (27.9) | 64,541 (20.9) |
| Chronic sinusitis, n (%) | 69,055 (17.4) | 22,905 (26.5) | 46,150 (14.9) |
| **Healthcare utilization** |  |  |  |
| Asthma ED or UC visit, n (%) | 93,856 (23.7) | 31,355 (36.3) | 62,501 (20.2) |
| **Laboratory** |  |  |  |
| Eosinophil counts (elevated), n (%) | 134,474 (33.9) | 32,825 (38.0) | 101,649 (32.8) |

Cases also had a higher prevalence of comorbid atopic conditions, including allergic rhinitis (27.9% vs 20.9%) and chronic sinusitis (26.5% vs 14.9%), as well as greater prior healthcare utilization, reflected by a higher proportion with asthma-related emergency department or urgent care visits (36.3% vs 20.2%). Elevated eosinophil counts were more common among cases than controls (38.0% vs 32.8%), consistent with greater inflammatory disease burden.

### Selected Feature Sets Across Dimensionality-Reduction Strategies

Feature-selection strategies yielded overlapping but non-identical subsets of variables, with attribution- and performance-based methods selecting features based on model-derived importance metrics, while the clinically guided subset emphasized interpretability and implementation feasibility (Table E1). These selected feature sets formed the basis for all reduced-dimensional model evaluations.

### Discrimination Performance Under Fixed-Length Sequence Design

Under the fixed-length sequence design, reduced-feature models demonstrated discrimination performance comparable to the full-feature reference model despite substantial reductions in input dimensionality (Table 3). Performance was evaluated on held-out MSAs in each fold and averaged across all six folds. The reference model achieved a mean AUROC (±SD) of 0.870 (±0.007) and an AUPRC of 0.718 (±0.034), with only modest differences across reduced-feature configurations.

**Table 3.** Discrimination Performance and Computational Efficiency Under Fixed-Length Sequence Design.

| Method | Number of | AUROC | AUPRC | Total Training | Inference Time |
| --- | --- | --- | --- | --- | --- |
| All features (reference) | 80 | $0.870 \pm 0.007$ | $0.718 \pm 0.034$ | $4.97 \pm 0.26$ | 292.73 |
| IG | 10 | $0.843 \pm 0.006$ | $0.664 \pm 0.035$ | $4.97 \pm 0.25$ | 292.65 |
| IG | 20 | $0.855 \pm 0.005$ | $0.687 \pm 0.031$ | $4.96 \pm 0.23$ | 291.99 |
| IG | 40 | $0.864 \pm 0.006$ | $0.698 \pm 0.028$ | $5.02 \pm 0.23$ | 295.39 |
| AUC | 10 | $0.829 \pm 0.004$ | $0.632 \pm 0.034$ | $4.96 \pm 0.27$ | 270.40 |
| AUC | 20 | $0.839 \pm 0.005$ | $0.670 \pm 0.020$ | $4.96 \pm 0.26$ | 291.99 |
| AUC | 40 | $0.846 \pm 0.006$ | $0.668 \pm 0.033$ | $5.02 \pm 0.31$ | 296.20 |
| Clinical | 20 | $0.836 \pm 0.005$ | $0.646 \pm 0.034$ | $4.90 \pm 0.29$ | 289.16 |

All results were obtained under a fixed-length sequence design with a maximum sequence length of 512, defined as the maximum number of structured EHR input elements included in a patient’s longitudinal history after chronological ordering. Batch size was fixed at 32 to ensure fair comparison across models. Values are reported as mean (standard deviation) across six cross-validation folds. The reference model uses the complete 80-feature input space without feature selection. AUROC indicates area under the receiver operating characteristic curve; AUPRC indicates area under the precision–recall curve. Total training time reflects end-to-end model training time on a single GPU-equivalent environment. Inference time represents the average latency required to generate a single prediction during inference.

Among Integrated Gradients–selected models, AUROC values ranged from 0.843 to 0.864 across 10-, 20-, and 40-feature configurations, with corresponding AUPRC values between 0.664 and 0.698 (Table 3). Discrimination differences remained modest despite substantial reductions in feature count.

Models using univariate AUROC-based selection showed similar patterns, although discrimination was consistently slightly lower than that of Integrated Gradients–based models at comparable feature counts.

The clinically guided 20-feature model achieved an AUROC of 0.836 (±0.005) and an AUPRC of 0.646 (±0.034), retaining most of the discriminatory performance of higher-dimensional models despite its more parsimonious feature set.

Total training time and inference latency were similar across models under the fixed-length sequence design, indicating that reductions in feature dimensionality did not materially affect computational performance in this setting (Table 3).

### Impact of Maximum Sequence-Length Truncation on Runtime and Discrimination

Sequence-length truncation was associated with substantial reductions in computational cost while preserving most discriminatory performance across a wide range of truncation thresholds (Table 4). Using the Integrated Gradients–selected 20-feature model as a representative configuration, setting the maximum sequence length at the 98th percentile of the empirical sequence-length distribution reduced total training time by approximately 38% relative to the fixed-length reference configuration, with minimal change in AUROC.

**Table 4.** Impact of Maximum Sequence Length on Discrimination and Computational Efficiency.

| Method | Number of Features | Maximum sequence length (max_len) | AUROC | Total Training Time (h) | Throughput (records/s) | $\Delta$ Training Time (%) | $\Delta$ AUROC |
| --- | --- | --- | --- | --- | --- | --- | --- |
| All features (reference) | 80 | 512 | $0.870 \pm 0.007$ | $4.97 \pm 0.26$ | $3.68 \pm 1.16$ | NA | NA |
| IG | 20 | 512 | $0.855 \pm 0.005$ | $4.96 \pm 0.23$ | $3.75 \pm 1.16$ | -0.1% | -0.015 |
| IG | 20 | p98 | $0.854 \pm 0.005$ | $3.08 \pm 0.17$ | $6.05 \pm 1.93$ | -38.0% | -0.016 |
| IG | 20 | p95 | $0.854 \pm 0.005$ | $2.23 \pm 0.12$ | $8.36 \pm 2.65$ | -55.2% | -0.016 |
| IG | 20 | p90 | $0.852 \pm 0.005$ | $1.70 \pm 0.09$ | $10.98 \pm 3.45$ | -65.9% | -0.018 |
| IG | 20 | p85 | $0.850 \pm 0.005$ | $1.47 \pm 0.08$ | $12.63 \pm 3.96$ | -70.3% | -0.020 |
| IG | 20 | p80 | $0.848 \pm 0.005$ | $1.32 \pm 0.07$ | $14.08 \pm 4.45$ | -73.4% | -0.022 |
| IG | 20 | p75 | $0.845 \pm 0.005$ | $1.13 \pm 0.06$ | $16.43 \pm 5.15$ | -77.2% | -0.025 |
| IG | 20 | p70 | $0.842 \pm 0.005$ | $1.01 \pm 0.06$ | $18.41 \pm 5.84$ | -79.6% | -0.028 |
| IG | 20 | p65 | $0.839 \pm 0.005$ | $0.94 \pm 0.06$ | $19.78 \pm 6.37$ | -81.0% | -0.031 |
| IG | 20 | p60 | $0.835 \pm 0.005$ | $0.84 \pm 0.04$ | $22.30 \pm 6.99$ | -83.2% | -0.035 |
| IG | 20 | p55 | $0.830 \pm 0.005$ | $0.77 \pm 0.04$ | $24.07 \pm 7.53$ | -84.4% | -0.040 |
| IG | 20 | p50 | $0.826 \pm 0.005$ | $0.73 \pm 0.04$ | $25.49 \pm 8.03$ | -85.3% | -0.044 |

Results are shown for the Integrated Gradients–selected model using 20 features; feature-selection method and feature count were held constant while maximum sequence length (max\_len) was varied. All models were trained using a fixed batch size of 32. Values are reported as mean (standard deviation) across six cross-validation folds. Maximum sequence length specifies the allowed number of structured EHR input elements per patient after chronological ordering. A value of 512 indicates the reference configuration, while values of pXX denote settings derived from the XXth percentile of the empirical sequence-length distribution. AUROC indicates area under the receiver operating characteristic curve. Total training time reflects end-to-end model training time (in hours) on a single GPU-equivalent environment. Throughput represents the average number of patient records processed per second during training. $\Delta$ Training Time (%) and $\Delta$ AUROC indicate differences relative to the reference configuration (max\_len = 512).

Progressively smaller maximum sequence lengths yielded larger gains in computational efficiency. Maximum sequence lengths corresponding to the 80th and 75th percentiles reduced training time by more than 70% compared with the reference configuration, while AUROC declines remained modest across cross-validation folds. In contrast, maximum sequence lengths below the 65th percentile were associated with more pronounced degradation in discrimination, suggesting diminishing returns when the allowed sequence length becomes overly restrictive (Table 4).

These relationships are illustrated in Figure 2, where training time decreased monotonically as maximum sequence length was reduced, whereas AUROC remained largely stable across a wide range of truncation thresholds, with only modest declines observed at the most restrictive settings.

Figure 3 shows the empirical distribution of sequence lengths, which was highly right-skewed, with most sequences relatively short and a small proportion substantially longer. The median sequence length was approximately 67, with the 80th, 90th, 95th, and 98th percentiles corresponding to sequence lengths of approximately 133, 186, 243, and 328, respectively.

### Calibration and Risk Re-Stratification Across Feature Sets

Calibration and risk stratification analyses evaluated the effects of feature dimensionality on predicted risk distributions, patient-level risk assignment, and high-risk identification (Figure 4).

**Figure 4.** Effect of feature dimensionality on calibration, risk stratification and event capture. Multi-panel figure illustrating how dimensionality reduction influences calibration, observation-level risk assignment, high-risk identification, and event capture across models. (**A**) **Calibration across the full risk distribution (upper left)** Calibration curves comparing predicted and observed risk for the full-feature reference model and reduced-feature models using shared risk bins. Risk bin boundaries were defined as quintiles (20% intervals) of predicted risk from the full-feature reference model and then applied unchanged to all reduced-feature models. In higher-risk bins, reduced-feature models exhibited lower predicted and observed risks than the reference model, consistent with attenuation of high-risk stratification with increasing dimensionality reduction. **(B)** Observation-level comparison of predicted risk relative to the full-feature reference model (upper right) Scatter plot comparing predicted risks from reduced-feature models against predictions from the full-feature reference model. Each point represents an individual observation, and the diagonal line indicates equality between models. Systematic deviation below the identity line at higher reference-model risk values indicates a narrower range of predicted risks assigned by reduced-feature models at the upper end of the risk spectrum. **(C)** Number of observations exceeding a reference-defined high-risk threshold (lower left) A fixed predicted-risk threshold was derived from the full-feature reference model to classify 20% of observations as high risk. Applying this same threshold to reduced-feature models, the number and percentage of observations classified as high risk are shown for each model configuration. The number and percentage of observations classified as high risk were similar across models, with only modest differences between the full-feature reference model and reduced-feature configurations. **(D)** Event capture at a fixed high-risk threshold (lower right) Number and percentage of observed asthma exacerbation events captured among observations classified as high risk using the same fixed threshold applied in Panel C. Event capture was similar across models, with only modest differences between the full-feature reference model and reduced-feature configurations. Sensitivity analyses examining broader risk-selection thresholds are shown in Figure E2.

Using shared risk bins defined by quintiles of predicted risk from the full-feature reference model, calibration plots demonstrated systematic differences between the reference and reduced-feature models in higher-risk strata (Figure 4A). Across reduced-feature configurations, mean predicted risk and observed event rates were lower in the highest-risk quintiles, indicating attenuation of risk estimates with increasing dimensionality reduction. Differences between models were minimal in lower-risk groups.

Patient-level comparisons further illustrated differences in risk assignment (Figure 4B). This analysis was restricted to patients with predicted risk ≥ 0.8 in the reference model and is shown for three representative 20-feature configurations (IG-20, AUC-20, and Clinical-20), based on a random sample of 1,000 patients per model. Across all reduced-feature models, predicted risks were systematically lower than those of the reference model, as indicated by deviation below the identity line. The degree of attenuation varied across feature-selection strategies, with IG-20 showing closer alignment and AUC-20 demonstrating greater downward deviation.

To assess the operational implications of these differences, a fixed high-risk threshold defined by the reference model (top 20% of patients) was applied across all models (Figure 4C). The proportion of patients classified as high risk remained close to 20% across configurations. For example, IG-10 and IG-20 identified slightly fewer high-risk patients (19.2% and 19.4%, respectively), whereas IG-40 identified a proportion similar to the reference model.

Using the same threshold, the proportion of observed asthma exacerbation events captured among high-risk patients was also similar across models (Figure 4D). IG-40 and IG-20 captured proportions of events most comparable to the reference model, with only modest differences observed across reduced-feature configurations.

### Sensitivity Analysis

In sensitivity analyses, discrimination performance was highly consistent across batch-size configurations (Table E2A). Mean AUROC values increased modestly with larger batch sizes, from 0.8479 at batch size 32 to 0.8495 at batch size 128, with differences ≤0.002 across folds.

Computational efficiency improved with increasing batch size. Average total training time decreased from 1.32 hours at batch size 32 to 1.16 hours at batch size 128, corresponding to an approximately 12% reduction in runtime (Table E2B).

In a separate sensitivity analysis, cumulative event capture was evaluated across a range of risk-selection thresholds (Supplementary Figure E2). Differences in event concentration across models were minimal at the operational threshold used in the main analysis (Figure 4D) and became more apparent only when broader fractions of patients (approximately 25–35%) were considered. Even under this worst-case ranking-based scenario, differences in cumulative event capture across models remained modest.

## DISCUSSION

In this study, we provide a systematic evaluation of input representation design in transformer-based clinical risk prediction using longitudinal EHR data. Across multiple feature-selection strategies and sequence-length configurations, we demonstrate that substantially more parsimonious input representations can preserve most discriminatory performance while markedly improving computational efficiency. Importantly, our findings show that reductions in feature dimensionality and sequence length alter predicted risk distributions, particularly at the upper tail, without materially affecting clinically relevant risk stratification or event capture at operational decision thresholds. These results highlight a critical distinction between global performance metrics and deployment-relevant behavior. By quantifying trade-offs between input complexity, computational burden, and clinically meaningful performance, this study provides practical guidance for scalable and deployable transformer-based models in real-world healthcare settings.

Longitudinal EHR-based prediction models are frequently constructed using large numbers of heterogeneous features and extended temporal histories in an effort to maximize predictive signal. [2, 4, 7] However, this approach introduces important practical challenges that are often underemphasized in model development. High-dimensional feature spaces increase data engineering complexity and may reduce portability across sites and workflows, while long input sequences substantially increase computational cost for transformer-based architectures. [8, 9, 22] Our findings suggest that much of this input complexity may be unnecessary for achieving strong and clinically meaningful predictive performance. Reduced feature sets and shorter sequences preserved discrimination and threshold-based risk stratification while substantially improving computational efficiency, supporting more parsimonious input design as a viable strategy for real-world implementation.

Prior studies examining dimensionality reduction in deep learning models for longitudinal EHR data have largely emphasized optimization-oriented metrics such as training loss, accuracy, and discrimination.[3, 10, 14, 15] In contrast, our study evaluated calibration, patient-level risk assignment, threshold-based risk stratification, and event capture under controlled comparisons of both feature dimensionality and sequence length. This integrated framework provides practical insight into how input representation choices influence both predictive performance and deployment-relevant behavior in transformer-based EHR models.

Figure 4 highlights an important distinction between distribution-level calibration and threshold-based clinical performance. [14, 16] Reduced-feature models exhibited attenuation of predicted risk and modest calibration differences at the upper end of the risk spectrum, particularly within the highest-risk group (Figure 4A–B). Despite these differences, performance at clinically relevant decision thresholds remained largely unchanged. When a fixed threshold identifying the top 20% of patients as high risk was applied, reduced-feature models identified a similar proportion of high-risk patients and captured a comparable fraction of observed asthma exacerbation events relative to the full-feature reference model (Figure 4C–D). Across feature-selection strategies, IG-based models (IG-40 and IG-20) showed performance most closely aligned with the reference model, whereas more aggressive reduction strategies demonstrated slightly greater attenuation. These findings suggest that dimensionality reduction introduces modest attenuation in calibration at the extreme upper tail of predicted risk, but with limited practical impact on threshold-based patient identification and event capture.

Although this study was not designed to evaluate clinical interventions, the observed differences in risk stratification across model configurations have important implications for how prediction models are operationalized in clinical settings. In practice, such models are typically deployed using fixed decision thresholds to identify patients for enhanced monitoring or preventive action. [9, 16]

Reduced-feature models, particularly when combined with moderate maximum sequence length designs, may offer a favorable balance between computational efficiency, data availability, and clinical performance in settings with constrained resources or frequent retraining requirements. Conversely, higher-dimensional models with longer effective input histories may better differentiate patients at the extreme upper end of the risk distribution, albeit with greater computational and operational complexity. These considerations are particularly relevant for health systems seeking to operationalize deep learning models under real-world constraints of data availability, computational resources, and model maintenance. [8, 9]

Longitudinal EHR data are characterized by highly skewed distributions of observed sequence length, with a small proportion of patients contributing disproportionately long histories. [2] By empirically characterizing this distribution and evaluating alternative maximum sequence length settings, we demonstrate that reducing the allowed input length yields substantial gains in computational efficiency. Moderate maximum sequence length settings reduced training time by more than half while preserving discrimination, whereas more restrictive maximum sequence lengths resulted in diminishing returns and larger performance degradation. Importantly, sequence-length selection was guided by empirical data characteristics rather than arbitrary architectural constraints, enhancing the practical relevance of the modeling framework.

The observed effects of dimensionality reduction and sequence-length design are likely dependent on the prediction task and the underlying data. Reduced-feature models may have less capacity to distinguish patients at the extremes of risk when variables encoding healthcare utilization intensity or complex nonlinear relationships are excluded. Similarly, the impact of sequence-length truncation will depend on the temporal structure of the prediction task and the distribution of sequence lengths of the underlying data. Accordingly, model design choices should be evaluated within the context of the specific clinical application.

Sequence-length truncation may have important implications for preserving longitudinal clinical information. Because sequence truncation removed the earlier portion of longer longitudinal EHR sequences, historical clinical information may have been excluded for patients with more extensive clinical histories. Potentially affected information could include historical exacerbations, prior medication exposures, treatment changes, or patterns of healthcare utilization that may provide important context regarding long-term disease severity. Consequently, patients with longer or more complex longitudinal histories may be more susceptible to information loss than patients with shorter histories. Although overall model discrimination remained stable across moderate sequence-length reductions, these aggregate performance metrics do not exclude the possibility that truncation could have a greater impact within specific patient subgroups. Accordingly, future implementation studies should evaluate model performance beyond population-level discrimination metrics, as aggregate performance measures may mask differences among patients with longer or more complex longitudinal histories. Subgroup validation stratified by historical sequence length or healthcare utilization may help determine whether sequence truncation disproportionately affects these patients before clinical deployment.

This study has several limitations that warrant consideration. First, we evaluated a limited set of predefined feature-selection strategies, including Integrated Gradients, univariate AUROC ranking, and a clinically guided feature subset. Other approaches, such as adaptive or dynamically learned feature selection methods, may further optimize the trade-off between computational efficiency and predictive performance. Second, all analyses were conducted within a single integrated healthcare system. Although this setting enables consistent longitudinal data capture and outcome ascertainment, generalizability to other healthcare systems may be limited. Third, computational evaluations were performed under controlled experimental conditions with fixed model architectures and training procedures. Real-world deployment may introduce additional constraints related to data latency, missingness, evolving feature availability, and retraining frequency that were not explicitly modeled in this study. In addition, we did not characterize the specific types of longitudinal clinical information removed by truncation or evaluate whether truncation differentially affected patients with longer or more complex disease histories.

In summary, transformer-based clinical risk prediction models can achieve strong performance using substantially more parsimonious input representations than is commonly assumed. Although reductions in feature dimensionality and sequence length introduce modest changes in calibration and risk distribution, these effects have limited impact on clinically relevant risk stratification and event capture at operational decision thresholds. These findings suggest that input complexity can be strategically evaluated and reduced to improve computational efficiency while maintaining strong overall predictive performance.

### Contributors

WC conceived the study with input from all co-authors and led the study design. Data extraction, preparation and analysis were performed by BZ. All authors contributed to interpretation of the results. WC drafted the manuscript, and all authors critically revised the manuscript for important intellectual content and approved the final version for submission. WC led funding acquisition for the study. WC, BZ and FX had access to the data. WC takes responsibility for the integrity of the data and the accuracy of the data analysis.

## Abbreviations

AAE: Acute asthma exacerbation
AUC: Area under the curve
AUPRC: Area under the precision–recall curve
AUROC: Area under the receiver operating characteristic curve
ED: Emergency department
EHR: Electronic health record
IG: Integrated Gradients
ICS: Inhaled corticosteroid
KPSC: Kaiser Permanente Southern California
LABA: Long-acting beta-agonist
max_len: Maximum sequence length
ML: Machine learning
MSA: Medical Service Area
PR: Precision–recall
ROC: Receiver operating characteristic
SABA: Short-acting beta-agonist
SD: Standard deviation
UC: Urgent care
XAI: Explainable artificial intelligence

## Acknowledgements

The authors thank the patients of Kaiser Permanente for helping to improve care through the use of information collected through our electronic health record systems. The authors also thank Ms. Sole Cardoso for her assistance with manuscript formatting and submission.

## Data sharing statement

Anonymized data that support the findings of this study may be made available from the investigative team in the following conditions: 1) agreement to collaborate with the study team on all publications, 2) provision of external funding for administrative and investigator time necessary for this collaboration, 3) demonstration that the external investigative team is qualified and has documented evidence of training for human subjects protections, and 4) agreement to abide by the terms outlined in data use agreements between institutions.

